# COVID-19 pandemic: A Hill type mathematical model predicts the US death number and the reopening date

**DOI:** 10.1101/2020.04.12.20062893

**Authors:** Yasser Aboelkassem

## Abstract

A mathematical model that can be used to estimate the total number of cases and deaths due to COVID-19 pandemic is presented in this study. The parameters and the associated uncertainty in the model are optimized and quantified using various reported data sets reported from different countries. The results suggest that, by the mid of June or early July 2020, the outbreak will strongly decay and the US will have about 800K confirmed cases and less than 50K deaths.

**Significance Statement:** This study presents a mathematical model that can be used to estimate the total number of cases and deaths in the US due to COVID-19. The model forecasting about < 800*K* cases and ~< 50*K* total numbers of deaths in the United States. The results suggest that late May or early June, 2020 is probably a good time to end the shutdown order and reopen the country for the daily routine business.

---

**C**oronavirus disease 2019 (COVID-19) pandemic is now a major worldwide life threat. On December 2019, the outbreak started in China and has rapidly spread to the entire world. The new COVID-19 outbreak is currently under investigations by many expert researchers around the world, trying to forecast its impact on our health system and framing questions for pandemic prevention (1–3)

As of April 12 2020, there have been more than 1.8M confirmed cases and more than 100K deaths worldwide. In the US, infection has started to rapidly spread to all over the states with about 8.5K deaths so far. Based on reported cases and death rate per day, probabilistic models predicted that there would be a hundred thousand if not millions of deaths globally (4, 5).

Although our current understanding of COVID-19 infection dynamics is still very limited. Yet, mathematical modeling, uncertainty quantification, and computational data analysis can be used to forecast an accurate measure of disease spread. These techniques can be used to forecast and number of mortality across countries during the time-course of the epidemic.

In this study, a Hill-type mathematical model is proposed to analyze the reported data from countries, namely the USA, Italy, Spain, and China. The model was then used to predict the total mortality number in these countries and forecast a safe reopening date in the US.

## Methods

Consider analyzing the COVID-19 data, including the time-tracking of the total number of confirmed cases and the number of deaths reported from the USA, Italy, Spain, and China. The modeling approach is divided into three major steps: (i) propose a Hill type mathematical dynamic model; (ii) parameter optimization; and (iii) predicting of the total numbers of cases and deaths, the death rate in the US and forecast a safe date of the re-open in the US. The details of each step are given below.

### Mathematical Model

The Hill equation ((6)) is frequently used to study the kinetics of reactions that exhibit a sigmoidal behavior. The rate of many transporter-mediated processes can be analyzed by the Hill equation. The idea here is to use a modified version of the Hill equation to describe the spread dynamics of COVID-19 which can be seen as a transporter-mediated process.

This modeling approach assumes that, at a given day during the outbreak, there will be a certain percentage of infection spread (concentration) that can eventually activate new patients which results in new deaths (reaction rates). As days progress, people are expected to respect the social distance guideline and shelter-in-place order. Thus, the infection spread capability will be reduced and the dynamics of having new cases are expected to attain a steady state behavior (saturation) i.e., there will be no more large numbers of new cases. In summary, this epidemic process can be considered as a rate of a carrier-mediated transport process and can be modeled using a modified version of Hill equation.

Based on the above mentioned analogy, the proposed Hill-like equation can be written as

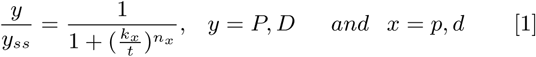

where, y represents the number of confirmed patients “P” cases or reported deaths “D”. The parameter *y*_*ss*_ refers to the steady-state (saturation) number in which there will be no more patients or deaths reported over time. The parameter t refers to the number of days. The parameter K (measured in days) is time at which the cases attains half of its projected maximum value. The exponent n refers to steepness “cooperativity” of this sigmoidal relationship which is commonly known as the Hill coefficient. The subscript x is used to alternate between patient and death modeling cases. It should be noted that equation (1) reveals three important parameters: *y*_*ss*_, K, and n, which must be optimized for prediction purpose.

### Parameter Optimization

The parameters involved in the above mathematical model are estimated using optimization technique which can be summarized as follows. In reported data ((7, 8)) from countries (USA, Italy, Spain, and China) with simultaneously tracking of the number of cases and deaths, we can use this data as an input and then optimize model parameters to obtain the best fit of the reported *PandD* curve. In other words, we minimize the RMS error between the calculated and the reported data of confirmed cases and deaths as:

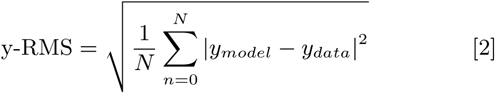

We have used a Matlab function *lsqnonlin* to find a set of model parameters which minimize RMS. This procedure will provide regression parameters (best-fit values) that define the shape and time for the curve. The fitted Hill-like equation will also follow a sigmoid shape at any value of n. In table (1), the optimized model parameters for the patients P cases of study and for each country under consideration are listed. Similarly, optimized parameters for the death D cases are listed in table (2).

**Table 1.**
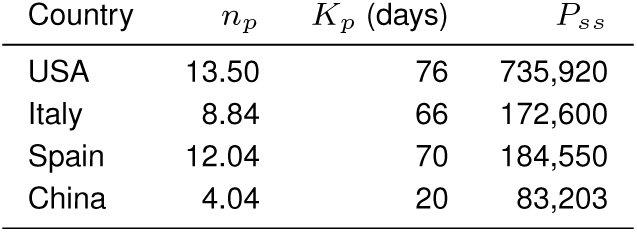
Model parameters which are used to predict the total number of confirmed cases in some countries.

**Table 2.** Model parameters which are used to predict the total number of deaths in some countries.

| Country | $n_d$ | $K_d$ (days) | $D_{ss}$ |
| --- | --- | --- | --- |
| USA | 17.12 | 81 | 41,285 |
| Italy | 9.92 | 71 | 24,135 |
| Spain | 14.08 | 73 | 20,005 |
| China | 3.67 | 26 | 3,427 |

### Predication

Once we have the optimal set of parameters as given in tables (1 and2), the number of patient cases P or the deaths D as a function of days during the course of this pandemic can be predicted. In other words, the model is simulated again, but using the optimized parameter sets for each country. This procedure will provide regression sigmoid curves which can be used to locate the COVID-19 saturation point in time for each country. This point can then be used to locate the total number of patients and deaths. Moreover, it can provide an estimated date of the society reopening to the normal daily activity.

## Results and Discussion

This study has generated results to estimate the total number of infected patients (cases) and deaths due to COVID-19 by day for the next few months. The results assume that social distancing guideline will continue to be respected throughout the epidemic interval.

In figure (1), the time course of the epidemic is shown for the USA, Italy, Spain, and China cases. The number of patients (P) over days is calculated using the optimized model parameters as explained in the previous section. The model was first validated against the data reported from China. The model prediction agrees well with the data from the four countries. The development of a number of cases as a function of days relationship shows a sigomidal behavior. The steady-state prediction of these curves suggest that, in the US we expect to have a total number of patients *P*_*ss*_ = < 800*K* cases. This number is lower in Italy (~180*K*) and Spain (~20*K*). No surprise, the model predicted an exact number for China, ~ 85*K*. The higher number patients in the US case of study can be explained by the steepness of the sigomidal response, where the Hill coefficient was estimated to be about *n*_*p*_ = ∼13.5 which is the highest among all countries in the world so far. The model also predicted the day (*K*_*p*_) that marks when we expect to reach exactly half of the saturation point of each country. In the US case, the results suggest a saturation of the epidemic by the end of May or early June 2020. This forecasting date can be used to estimate the end of the shutdown or lock-in-place orders.

**Fig. 1.**
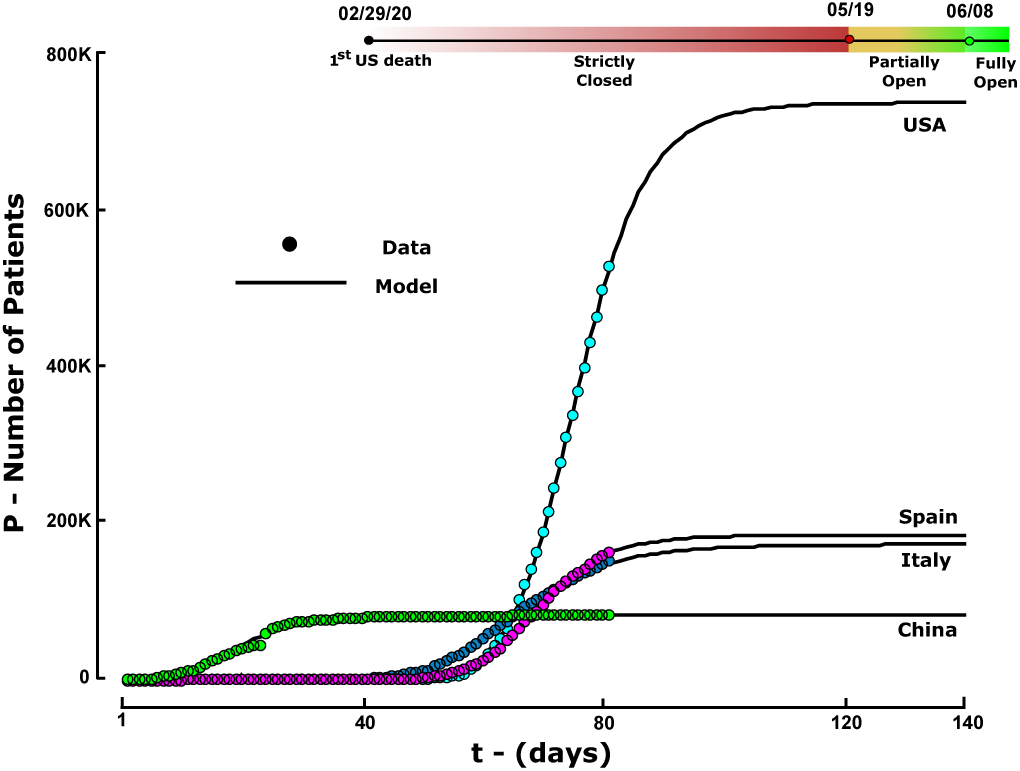
The time course of the epidemic is shown for the USA, Italy, Spain, and China cases. The number of patients (P) over days is calculated using the optimized model parameters. The model prediction agrees well with the data from the four countries. The model prediction of the expected timeline of the reopening date is shown on the top of the same figure.

Similarly, the number of deaths (D) over days is calculated using the same model and is shown in figure (2) for the same four countries. The model prediction agrees well with the available reported death data. The number of deaths as a function of days exhibits a sigomidal behavior. The steady-state of these curves suggest that, in the US we expect to have a total number of deaths *D*_*ss*_ = ∼50*K*. This number is expected to be lower in Italy (~25*K*), Spain (~22*K*) and China is about ~ 3.5*K*. The higher number deaths in the US are also explained by the steepness of the sigomidal death response curve, i.e., *n*_*D*_ = ∼17.12 which is the highest among all the simulated countries.

**Fig. 2.**
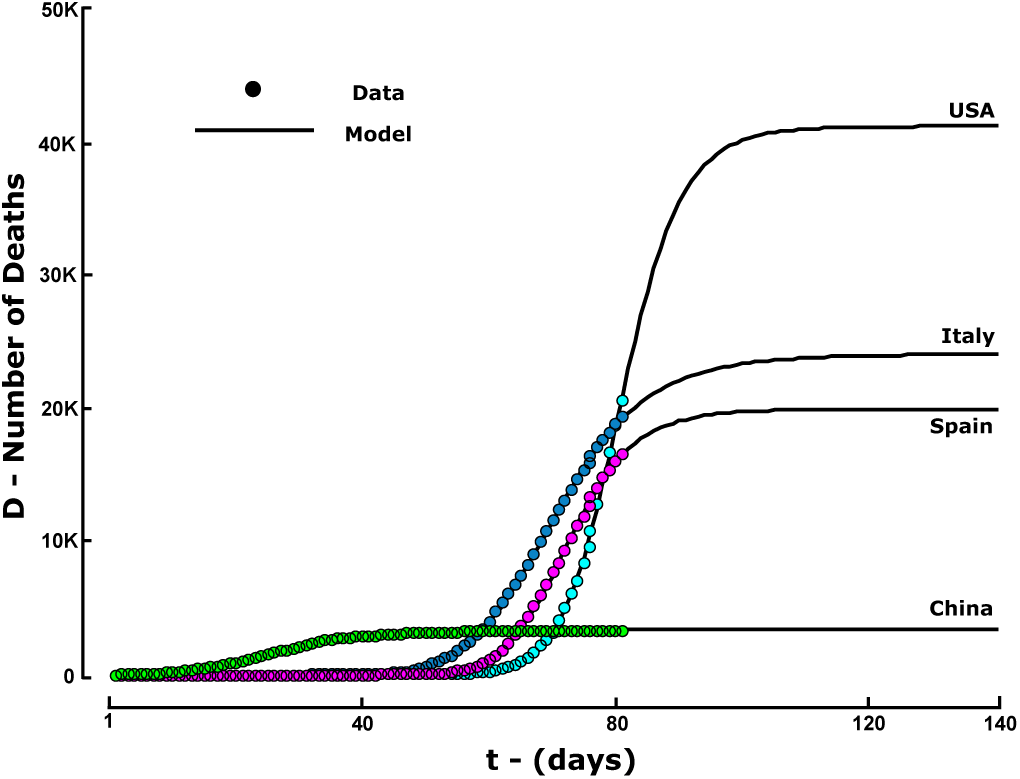
The number of deaths (D) as a function of time measured in days is calculated for the same four countries. The model prediction agrees well with the available reported death data.

In figure (3), the number of deaths per day is given for the same countries. The model suggests that the death rate exhibits a Gaussian or a bell-like distribution. The peak of each distribution occurs at day that is different from one another. For instance and according to the present analysis, the US death peak didn’t occur yet. This peak is expected to occur on April 15, 2020 leaving behind about (∼2.1*K*) deaths during this date. Similar results can be obtained for both Italy and Spain cases. The results suggested that the death rate has started to decline in both Italy and Spain.

**Fig. 3.**
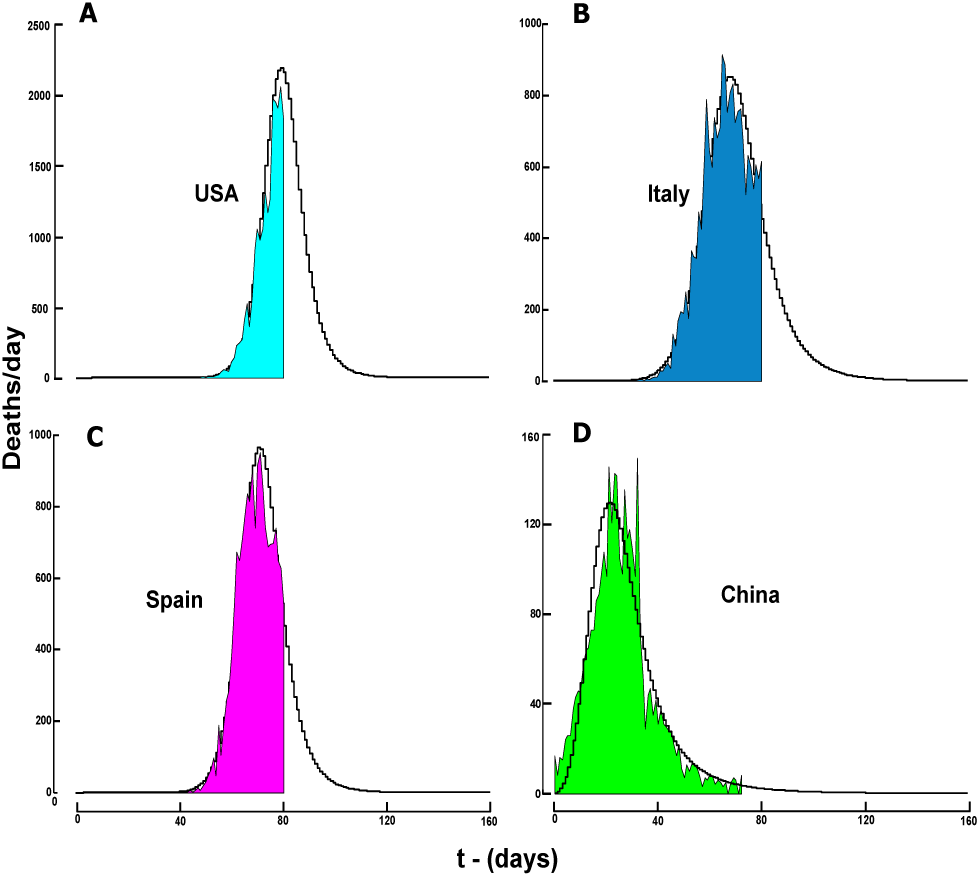
The number of deaths per day. The color shaded area presents the collected data up to date from USA, China, Italy, and Spain. The solid stair line presents the model prediction over the entire course of the epidemic.

In figure (4), the total number of deaths in the USA, China, Italy, and Spain represented by polar plots. The filled circles indicate the data collected. The solid spiral line refers to the model prediction parameterized by the number of days. The radius of the dotted circle refers to the total number of deaths. The model predicts a total death of about *D*_*ss*_ = 50*K* by the end of this pandemic. This is clearly shown from the spiral trajectory which attains a steady state value by mid May, 2020. In other words, the number of mortality in the US will stay bounded by a circle having a radius of ~ 50*K* deaths. Of course, as time progress this pandemic decays and assuming that all the necessary medical equipment (ICUs and Ventilators, etc..) becomes available, the total number of deaths in the US can be significantly reduced.

**Fig. 4.**
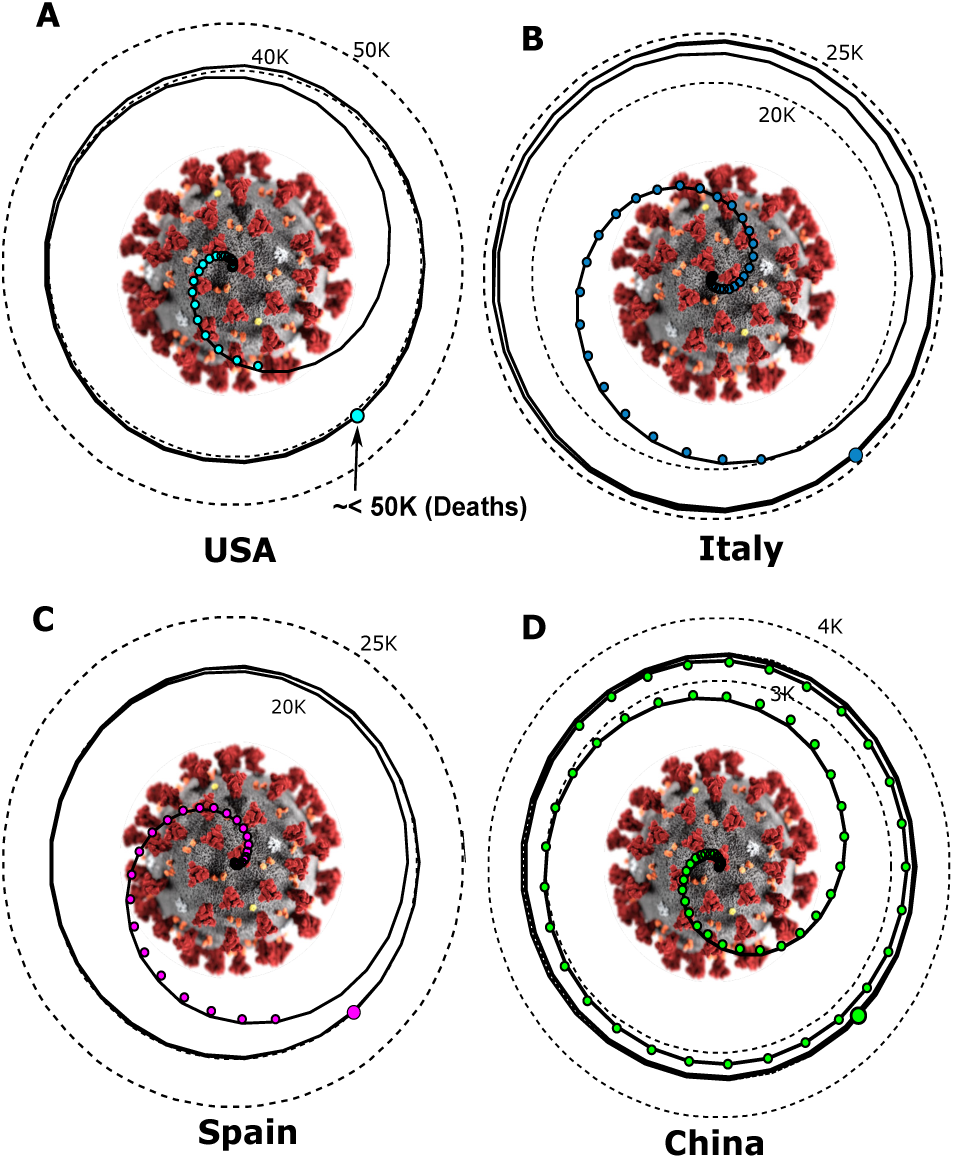
Prediction of the total number of deaths in the USA, China, Italy, and Spain represented by polar plots. The filled circles indicate the data collected. The solid spiral line refers to the model prediction parameterized by the number of days. The radius of the dotted circle refers to the total number of deaths. The virus illustration is created by the Centers for Disease Control and Prevention (CDC).

## Model Uncertainty and Numerical Algorithm

Uncertainty in the input data (patients and deaths) is considered when building this model using non-intrusive approach. Each input data point was sampled from a normal distribution *x*_*data*_ *∼ 𝒩*(*µ, σ*^2^) using Monte Carlo (MC) simulations. A mean (*µ*) value equal to the average data obtained from different sources and a variance of (*σ* = 0.25*µ*) was used. The Levenberg–Marquardt (LM) algorithm is used to solve non-linear least squares minimization problems (9).

## Data Availability

NA

## Acknowledgment

The author would like to thank Mr. Abdelrhman Aboelkassem for collecting and organizing the data used in this model.

